# General practice workforce retention strategies: an umbrella review

**DOI:** 10.1101/2025.01.21.25320715

**Authors:** L. Jefferson, S. Golder, A. Castro, E. Webster, H. Anderson, V. Dale

## Abstract

**Objective:** We summarise international evidence of interventions to support retention of the general practice workforce, mapping evidence by country, method and workforce group.

**Design:** Umbrella overview of reviews. We mapped evidence according to occupational group, intervention type and outcome to identify trends and gaps.

**Setting:** International primary care and general practice.

**Participants:** General practice workforce, with studies grouped by general practitioners, nurse practitioners and mixed health professional.

**Main outcome measures:** Workforce retention, including length of employment, stability rates, settlement rates, vacancy rates, unfilled positions, turnover numbers or rates, attrition rates, intentions to stay, leave and return.

**Results:** Of the 3449 articles identified through database searching and citation tracking, we included 12 reviews. There was a general lack of policy evaluations in some fields and low geographical spread beyond Australia and the US. While stronger evidence exists for interventions such as professional development and fostering supportive environments, mixed findings were reported on financial incentives and limited formal evaluations were found for some intervention types, and professional groups.

**Conclusion:** This overview of research underscores the need for more research evaluating interventions to support the general practice workforce, particularly across wider international settings.

## Introduction

Imbalance in healthcare workforce capacity is challenging general practice workforces internationally. In the UK, rising patient demand due to aging populations and backlogs of unmet need generated during the COVID-19 pandemic^1^ ^2^ has not been sufficiently met with workforce increases. General practitioner (GP) participation rates, measured as full time equivalents (FTE), are decreasing (from 0.80 to 0.69 median FTE since 2015)^3^ and around two thirds of GPs in England are considering leaving the profession within five years.^4–6^ Research has reported increases in burnout and mental health problems driven by rising patient demand,^1^ ^2^ toxic workplace cultures, and poor work-life balance, which have been directly linked to intentions to leave.^7^ ^8^ Across the NHS as a whole, record rates of sickness absence also impact participation rates – in August 2021 almost 2 million sickness days were lost across the NHS in one month alone, with 560,000 of these due to mental health problems.^9^

Questions exist around the future of the general practice model if trends continue.^10^ ^11^ Indeed, projected shortfalls of 1 in 4 GP posts (8,800 FTE GPs) and 1 in 2 general practice nurse posts are estimated by 2030/31 if trends continue.^12^ Practices are closing in some parts of the country, leaving patients without a local GP and resulting in increased workload for remaining practices; often in deprived areas where health needs are greatest.^13^ ^14^ Fewer GPs are now taking on roles as GP partners – 10% fewer over the past 5 years^11^ – fuelling practice closures and resulting in higher workloads and risk for remaining GP partners.^15^ ^11^ GPs also report increasing attractiveness of ‘portfolio roles’ that are associated with fewer clinical hours in order to manage challenging workloads.^8^ ^9^ ^16^

To solve workforce capacity issues, research and policy vary in their approach to either encourage recruitment, retention or return of the workers. In general practice, recruitment interventions dominate international research, particularly focused on underserved or rural areas.^17^ These strategies are an expensive, and long-term solution – at a cost of nearly £500,000^18^ and taking nearly ten years to train a fully-qualified GP in the UK. As a result, a shift has been seen towards retention strategies.^19^ ^20^ The Department of Health and Social Care now recognise ‘shaping and supporting the workforce’ as one of three Areas of Research Interest outlined by the UK Department of Health and Social Care.^21^ Developing strategies to support and retain this workforce is needed to meet the quadruple aim of healthcare, giving emphasis on provider wellbeing alongside previously recognised aims of improving patient experience, population health and reducing healthcare costs.^22^ This is welcome, not least because doctor wellbeing is important in its own right, but also since it effects the quality of patient care.^23^ ^24^

In 2023, the Royal College of General Practitioners stressed a need for a comprehensive review of existing retention initiatives to support future policy and research.^11^ Our umbrella review aims to meet this need through a summary of the international evidence of interventions to support retention of the general practice workforce. Specifically, we map evidence by country, method and workforce group to enable insights for future research and policy.

## Methods

Following the PRIOR reporting guideline for undertaking overviews of reviews^25^ and Cochrane guidelines for systematic reviews^26^ we searched databases (Medline, Embase, PsychInfo and Medrxiv) for reviews of general practice workforce retention conducted from 2010 onwards. To focus our review on empirical evidence evaluating interventions, we limited our searches to include only systematic reviews and scoping reviews, using the criteria below. Full searches are available in Appendix 1.

### Inclusion criteria

#### Type of participants

Studies in the general practice workplace setting were considered, including doctors, nurses and other primary care health professionals. Trainee groups were excluded. International terminology was considered, for example the use of ‘family practice’ in the USA. Owing to the large number of reviews in hospital care settings, only reviews with a focus on general practice or primary care settings were included.

#### Type of intervention

Interventions at either an individual, practice or system level were included.

#### Type of outcome measures

Measures of workforce retention, turnover or future intentions were included (for example, length of employment, stability rates, settlement rates, vacancy rates, unfilled positions, turnover numbers or rates, attrition rates, intention to stay/leave, intentions to return).

#### Types of study design

Review methodologies were included, including systematic reviews, meta-analyses, scoping reviews and reviews of reviews.

No language limits were placed on the search. Google Scholar was also searched, as were reference lists using backwards and forwards citation searching. Reviews of multiple health professional groups were excluded, owing to their tendency to not report outcomes separately for general practice workers. Reviews focusing only on incentivising rural practice retention were excluded since they addressed slightly different aims (retention to a location rather than broader workforce retention). These also tended to be based in countries with particular issues of rurality; such as Australia, Canada and USA.

### Selection of studies

We used the software tool Covidence^34^ to download all records and conduct screening, firstly of titles and abstracts, and later screening full texts of articles deemed potentially relevant. Both stages were undertaken by two independent reviewers (LJ, SG, AC, EW, HA or VD), with arbitration by a third reviewer where necessary. Excluded articles and reasons for exclusion were used to generate a flow diagram as recommended by the PRISMA statement.^35^

### Data extraction

Data extraction was performed by two reviewers (AC, LJ) using a pre-piloted data extraction form, with a 20% sample cross-checked and inter-rater reliability assessed. Information was extracted regarding country, number of included studies, proportion of studies with focus on rurality, search dates, health professional group, nature and direction of outcomes. Based on a framework previously used in reviews by the World Health Organisation^36^ and a Cochrane review,^37^

### Data synthesis

We use evidence mapping according to country, outcomes, professional groups and nature of intervention to outline evidence gaps. We categorised interventions as either: 1) Educational interventions, 2) Financial interventions, 3) Regulatory strategies or 4) Personal and professional support strategies.

## Results

### Search results

We identified 3449 articles through database searching and citation tracking. After removing duplicates, we identified 2484 articles that were screened as titles and abstracts. At this stage, we excluded 2232, leaving 252 for full text review. In total, 12 reviews^17^ ^38–48^ were identified that met our inclusion criteria (Figure 1).^49^

**Figure 1:**
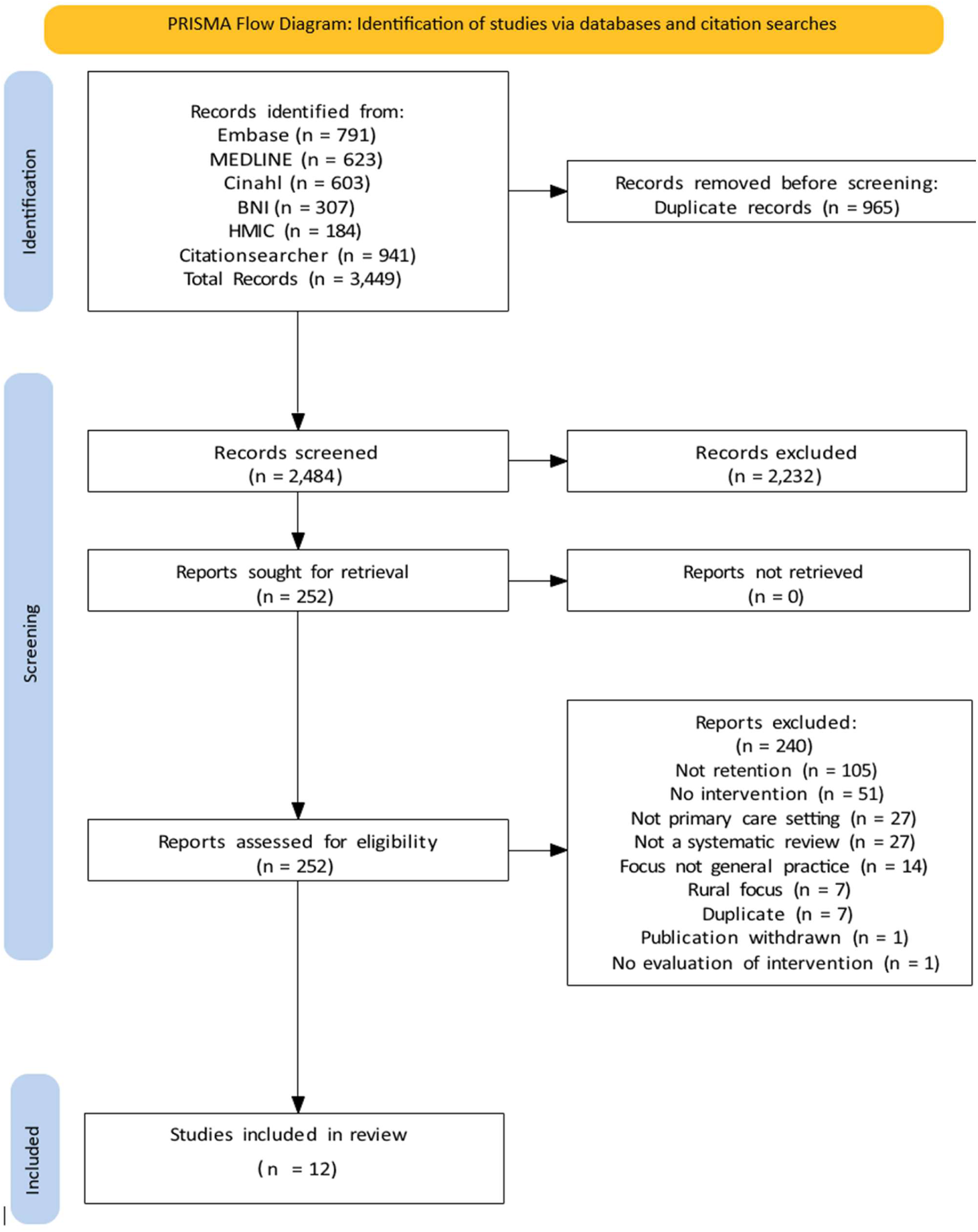
**PRISMA diagram**

### Study characteristics

Three reviews were focused on the nursing workforce in primary care settings,^39^ ^41^ ^48^ six were of GPs or primary care doctors,^17^ ^42^ ^44–47^ and three were of mixed professional groups in primary care.^38^ ^40^ ^43^ Studies included in the reviews were undertaken between 1974 and 2019; the most recent search date of included reviews was 2019.

Figure 2 demonstrates the geographical spread of searches of the included reviews; highlighting clear gaps where evidence has not yet been searched. Studies included in the reviews on this topic were predominantly located in Australia and USA (Table 1).

**Figure 2:**
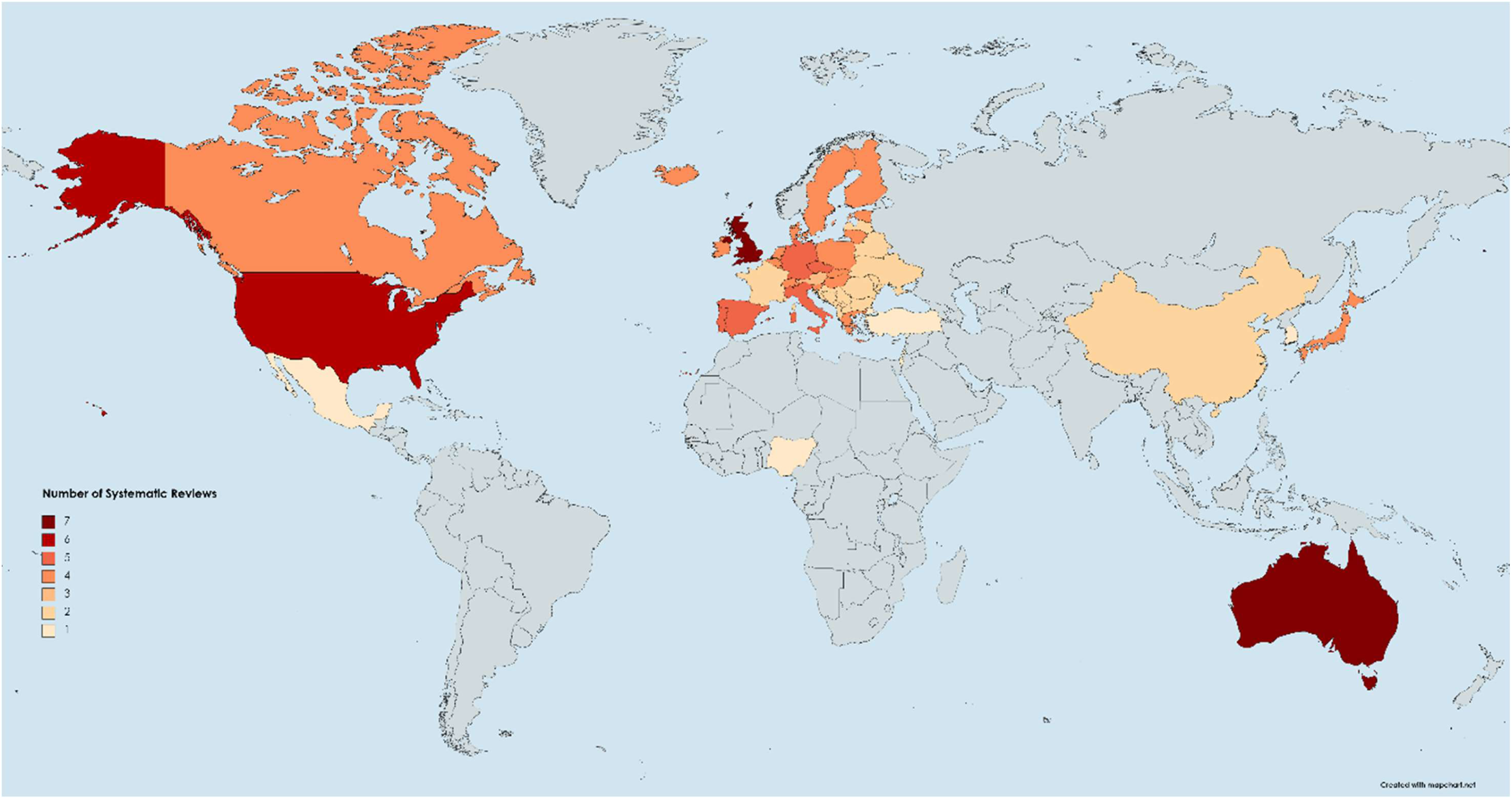
**Locations of search strategies of included reviews**

**Table 1:** Principal characteristics of reviews and findings according to intervention groups.

| Author | HCP group |  |  | Interventions |  |  |  |  | Total studies | Countries of relevant* included studies |
| --- | --- | --- | --- | --- | --- | --- | --- | --- | --- | --- |
|  | Nurse | GP | Mixed | Ed | Fin | Reg | PPS | Other |  |  |
| Walker 2020 |  |  |  |  |  |  |  |  | 12 | 1 AU |
| Kueakomoldej 2022 |  |  |  |  |  |  |  |  | 22 | 17 US |
| Lucas 2020 |  |  |  |  |  |  |  |  | 19 | 1 Brazil, 1 US, 1 AU, 1 Nigeria, 1 South Africa, 1 Finland |
| Marchland 2017 |  |  |  |  |  |  |  |  | 36 | 10 UK, 1 Lithuania, 1 AU, 1 Canada |
| Peckham 2016 |  |  |  |  |  |  |  |  | Unclear | Unclear |
| Mitchell 2018 |  |  |  |  |  |  |  |  | Unclear | Unclear |
| Verma 2016 |  |  |  |  |  |  |  |  | 51 | 8 US, 2 Canada, 2 AU, 1 Norway, 4 UK |
| Murray 2016 |  |  |  |  |  |  |  |  | 4 | 1 AU |
| Pettigrew 2016 |  |  |  |  |  |  |  |  | 5 | 1 UK |
| McCalman 2019 |  |  |  |  |  |  |  |  | 35 | 6 AU, 1 UK |
| Li 2019 |  |  |  |  |  |  |  |  | 36 | 7 China |
| Deroy 2019 |  |  |  |  |  |  |  |  | 26 | 6 AU, 1 NZ, 1 Mexico, 1 US |

**Abbreviations:** Interventions: Ed: Education, Reg: Regulation, Fin: Financial, PPS: Personal and professional support. Countries: UK: United Kingdom, US: United States, NZ: New Zealand, AU: Australia
\*Some reviews included wider workforce interventions not focused on retention. Figures represent retention studies.

### Evidence mapping

Our heatmaps highlight a general lack of evidence across the board. Research has focused more on doctors (GPs or family doctors) and mixed professional groups in included reviews, and reviews tend to include more evidence relating to interventions that sought to provide personal and professional support to individuals. Regulatory and educational interventions were less common – these tend to be used more for recruitment strategies. Some studies also referred to ‘new ways of working,’ which is an approach outlined in the 10 part plan developed by NHS England, Health Education England and key professional bodies (RCGP, BMA).^50^ Table 1 provides a summary of the included reviews, with mapping of the evidence by professional group, intervention type and location of included studies. Table 2 gives a heatmap by intervention type, and Table 3 by professional group.

**Table 2:**
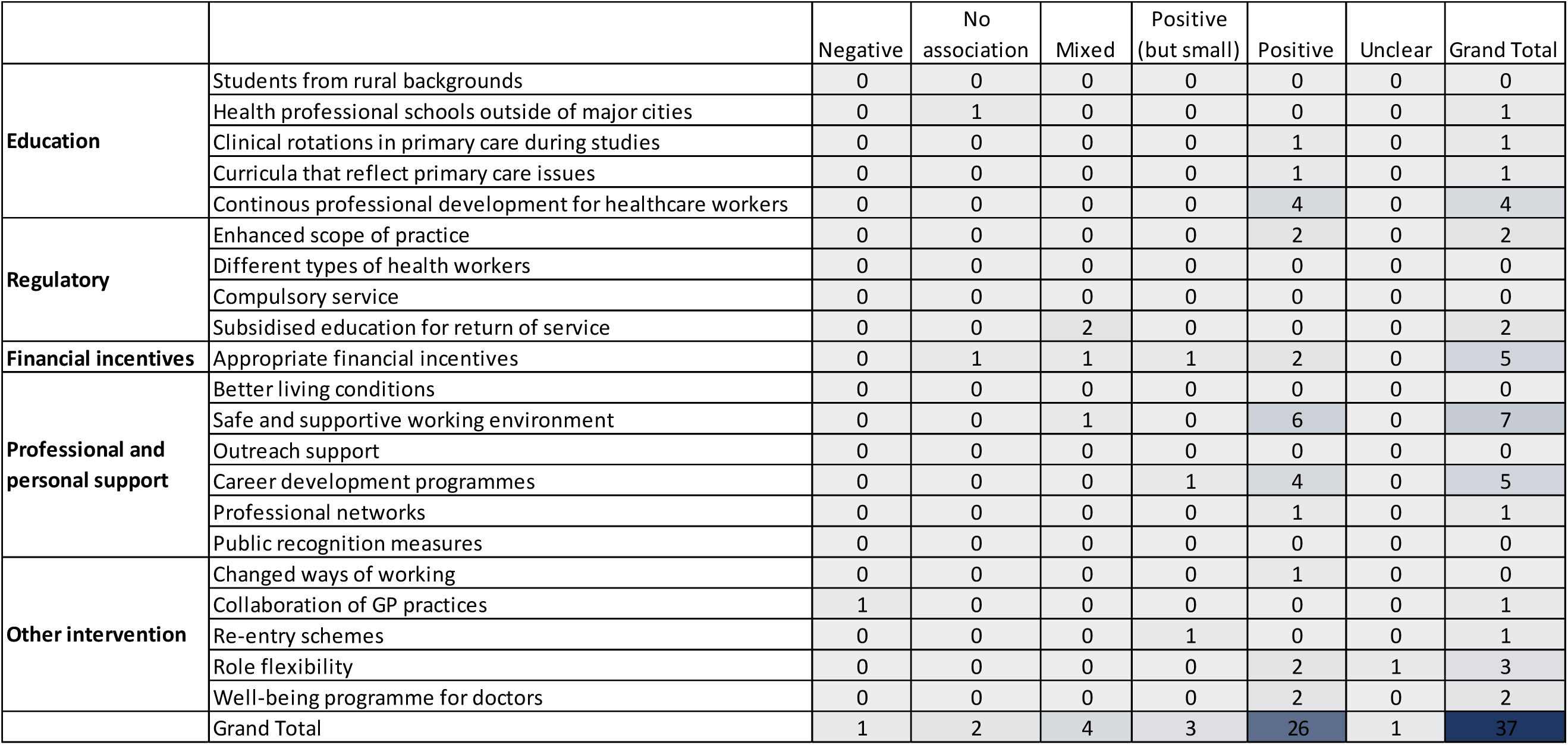
Heatmap of interventions by outcome, all reviews.

**Table 3:** Heatmap of interventions by outcome and occupational group.

| Occupational group | Intervention | Outcome |  |  |  |  |  | Grand Total |
| --- | --- | --- | --- | --- | --- | --- | --- | --- |
|  |  | Negative | No association | Mixed | Positive (but small) | Positive | Unclear |  |
| GPs | Students from rural backgrounds | 0 | 0 | 0 | 0 | 0 | 0 | 0 |
|  | Health professional schools outside of major cities | 0 | 1 | 0 | 0 | 0 | 0 | 1 |
|  | Clinical rotations in primary care during studies | 0 | 0 | 0 | 0 | 0 | 0 | 0 |
|  | Curricula that reflect primary care issues | 0 | 0 | 0 | 0 | 0 | 0 | 0 |
|  | Continuous professional development for healthcare workers | 0 | 0 | 0 | 0 | 0 | 0 | 0 |
|  | Enhanced scope of practice | 0 | 0 | 0 | 0 | 0 | 0 | 0 |
|  | Different types of health workers | 0 | 0 | 0 | 0 | 0 | 0 | 0 |
|  | Compulsory service | 0 | 0 | 0 | 0 | 0 | 0 | 0 |
|  | Subsidised education for return of service | 0 | 0 | 1 | 0 | 0 | 0 | 1 |
|  | Appropriate financial incentives | 0 | 1 | 1 | 1 | 0 | 0 | 3 |
|  | Better living conditions | 0 | 0 | 0 | 0 | 0 | 0 | 0 |
|  | Safe and supportive working environment | 0 | 0 | 0 | 0 | 2 | 0 | 2 |
|  | Outreach support | 0 | 0 | 0 | 0 | 0 | 0 | 0 |
|  | Career development programmes | 0 | 0 | 0 | 1 | 2 | 0 | 3 |
|  | Professional networks | 0 | 0 | 0 | 0 | 0 | 0 | 0 |
|  | Public recognition measures | 0 | 0 | 0 | 0 | 0 | 0 | 0 |
|  | Changed ways of working | 0 | 0 | 0 | 0 | 1 | 0 | 1 |
|  | Collaboration of GP practices | 1 | 0 | 0 | 0 | 0 | 0 | 1 |
|  | Re-entry schemes | 0 | 0 | 0 | 1 | 0 | 0 | 1 |
|  | Role flexibility | 0 | 0 | 0 | 0 | 2 | 1 | 3 |
|  | Well-being programme for doctors | 0 | 0 | 0 | 0 | 2 | 0 | 2 |
|  | Grand Total | 1 | 2 | 2 | 3 | 9 | 1 | 18 |
| Nurses | Students from rural backgrounds | 0 | 0 | 0 | 0 | 0 | 0 | 0 |
|  | Health professional schools outside of major cities | 0 | 0 | 0 | 0 | 0 | 0 | 0 |
|  | Clinical rotations in primary care during studies | 0 | 0 | 0 | 0 | 1 | 0 | 1 |
|  | Curricula that reflect primary care issues | 0 | 0 | 0 | 0 | 1 | 0 | 1 |
|  | Continuous professional development for healthcare workers | 0 | 0 | 0 | 0 | 1 | 0 | 1 |
|  | Enhanced scope of practice | 0 | 0 | 0 | 0 | 1 | 0 | 1 |
|  | Different types of health workers | 0 | 0 | 0 | 0 | 0 | 0 | 0 |
|  | Compulsory service | 0 | 0 | 0 | 0 | 0 | 0 | 0 |
|  | Subsidised education for return of service | 0 | 0 | 1 | 0 | 0 | 0 | 1 |
|  | Appropriate financial incentives | 0 | 0 | 0 | 0 | 1 | 0 | 1 |
|  | Better living conditions | 0 | 0 | 0 | 0 | 0 | 0 | 0 |
|  | Safe and supportive working environment | 0 | 0 | 1 | 0 | 2 | 0 | 3 |
|  | Outreach support | 0 | 0 | 0 | 0 | 0 | 0 | 0 |
|  | Career development programmes | 0 | 0 | 0 | 0 | 1 | 0 | 1 |
|  | Professional networks | 0 | 0 | 0 | 0 | 0 | 0 | 0 |
|  | Public recognition measures | 0 | 0 | 0 | 0 | 0 | 0 | 0 |
|  | Grand Total | 0 | 0 | 2 | 0 | 8 | 0 | 10 |
| Mixed professions | Students from rural backgrounds | 0 | 0 | 0 | 0 | 0 | 0 | 0 |
|  | Health professional schools outside of major cities | 0 | 0 | 0 | 0 | 0 | 0 | 0 |
|  | Clinical rotations in primary care during studies | 0 | 0 | 0 | 0 | 0 | 0 | 0 |
|  | Curricula that reflect primary care issues | 0 | 0 | 0 | 0 | 0 | 0 | 0 |
|  | Continuous professional development for healthcare workers | 0 | 0 | 0 | 0 | 3 | 0 | 3 |
|  | Enhanced scope of practice | 0 | 0 | 0 | 0 | 1 | 0 | 1 |
|  | Different types of health workers | 0 | 0 | 0 | 0 | 0 | 0 | 0 |
|  | Compulsory service | 0 | 0 | 0 | 0 | 0 | 0 | 0 |
|  | Subsidised education for return of service | 0 | 0 | 0 | 0 | 0 | 0 | 0 |
|  | Appropriate financial incentives | 0 | 0 | 0 | 0 | 2 | 0 | 2 |
|  | Better living conditions | 0 | 0 | 0 | 0 | 0 | 0 | 0 |
|  | Safe and supportive working environment | 0 | 0 | 0 | 0 | 2 | 0 | 2 |
|  | Outreach support | 0 | 0 | 0 | 0 | 0 | 0 | 0 |
|  | Career development programmes | 0 | 0 | 0 | 0 | 1 | 0 | 1 |
|  | Professional networks | 0 | 0 | 0 | 0 | 1 | 0 | 1 |
|  | Public recognition measures | 0 | 0 | 0 | 0 | 0 | 0 | 0 |
|  | Grand Total | 0 | 0 | 0 | 0 | 10 | 0 | 10 |

There is some evidence of a potential positive effect from studies that have explored interventions using continuous professional development, promoting a safe and supportive working environment and career development programmes.

For GP retention, there appears to be mixed evidence as to the potential effect of providing financial incentives. 4/10 interventions exploring nursing retention were categorised as promoting a safe and supportive working environment, while others were spread across different intervention types. In reviews of mixed primary care health professional groups, there is some evidence to suggest that continuous professional development for healthcare workers may have a positive effect on retention.

Only one review found a potentially *negative* effect of interventions when considering retention – Pettigrew^50^ explored the use of working at scale in UK general practice and reported potential negative impact on retention. However, this was based on the evidence from one qualitative study.

While retention was a focus for this review, this was often a secondary outcome in included studies. For example, policy interventions identified in Pettigrew’s review of working at scale tended to focus on patient outcome measures, rather than workforce outcomes. Harrington^52^ explored the use of mentoring in nurse practitioner programmes but no evaluations were identified, so this review was excluded from our search. Peckham^17^ and an updated review by Mitchell^44^ also did not find any formal evaluations of GP retention strategies, instead providing summaries of studies exploring factors associated with retention.

## Discussion

### Summary of findings

This research provides an overview of the scope and areas of focus from reviews addressing workforce retention in primary care settings. The included reviews span studies conducted between 1974 and 2019. There are notable gaps, both in terms of when the reviews were undertaken (most recently in 2019) and by geography, with lower coverage of countries outside Australia and the US. While there have been summaries of UK evidence,^17^ ^42^ ^44^ these reviews have not retrieved any experimental or quasi-experimental evaluations of interventions to support retention of general practice workforce.

Our evidence heatmaps revealed a general paucity of evidence across many intervention types and geographic regions, though stronger evidence exists in terms of interventions that seek to provide individual practitioners with personal and professional support. Interventions exploring continuous professional development, fostering safe working environments, and career development programs showed positive effects on retention. There appears to be mixed evidence as to the effectiveness of financial incentives for GPs, and evidence from one qualitative study^46^ suggests working at scale may produce negative associations with GP retention, though further research is now needed.

Across reviews, we found that workforce retention often featured as a secondary outcome, with many interventions primarily targeting patient outcomes or broader organisational goals.

Meanwhile, a review of nurse practitioner mentoring^52^ and reviews of GP retention^17^ ^44^ highlight a lack of formal evaluations, emphasising the need for more focused research in these areas.

### Implications for practice, policy, and future research

Further work is now needed to synthesize the evidence base since 2019, with a specific focus on retention of the primary care workforce. Historically, based on the most recent searches, gaps existed in evidence for regulatory and educational interventions. An updated review may highlight whether this gap has been filled, or whether more research is needed of retention-specific strategies across professional groups and global contexts.

### Strengths and limitations

This umbrella review provides a valuable insight into the existing evidence base, with this approach offering a high-level overview of the research gaps for interventions to support general practice workforce retention. This may help to prioritise future research efforts and policies to support future workforce retention. In addition to the geographical limitations of the searches of included review, there were also challenges in terms of how well evidence was reported in some studies. For example, it was common for reviews to report findings for recruitment and retention together, but often challenging to disentangle the evidence due to reporting.

### Conclusion

This research identifies significant gaps in geographical coverage of existing reviews in this field and underscores the needs for more targeted workforce retention research.

### Appendix 1: Search strategy

**Table 1:** Summary of inclusion and exclusion criteria.

| Inclusion | Exclusion |
| --- | --- |
| Primary care workforce. Using UK terminology such as general practitioners or international terminology such as 'family practice' or 'family physician.' | Reviews of medical specialities outside primary care (acute or hospital medicine), owing to their tendency to not report outcomes separately for primary care workers. |
| Interventions at either an individual, practice or system level. |  |
| Measures of workforce retention and intentions to leave practice were included (e.g.: length of employment, stability rates, settlement rates, vacancy rates, unfilled positions, turnover numbers or rates, attrition rates, intention to stay/leave, intentions to return). | Reviews not measuring retention (e.g. those solely exploring recruitment factors or patient outcomes). |
| Review methodologies (including systematic reviews, literature reviews or 'evidence syntheses', meta-analyses, scoping reviews and umbrella reviews). | Primary studies were excluded. Reviews not providing summaries of evaluations of interventions were excluded. |
| Published after 01.01.2010 | Published prior to 2010 |
| All languages |  |

## Database Search Strategies

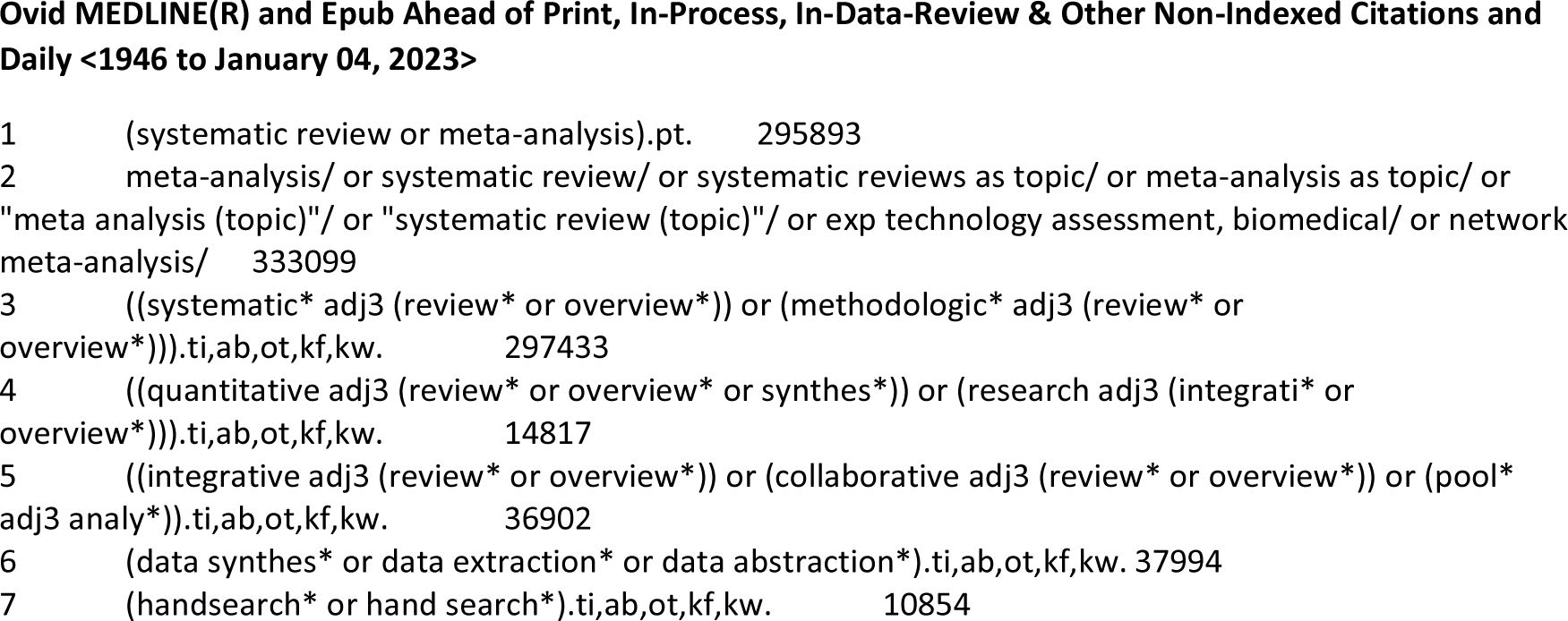
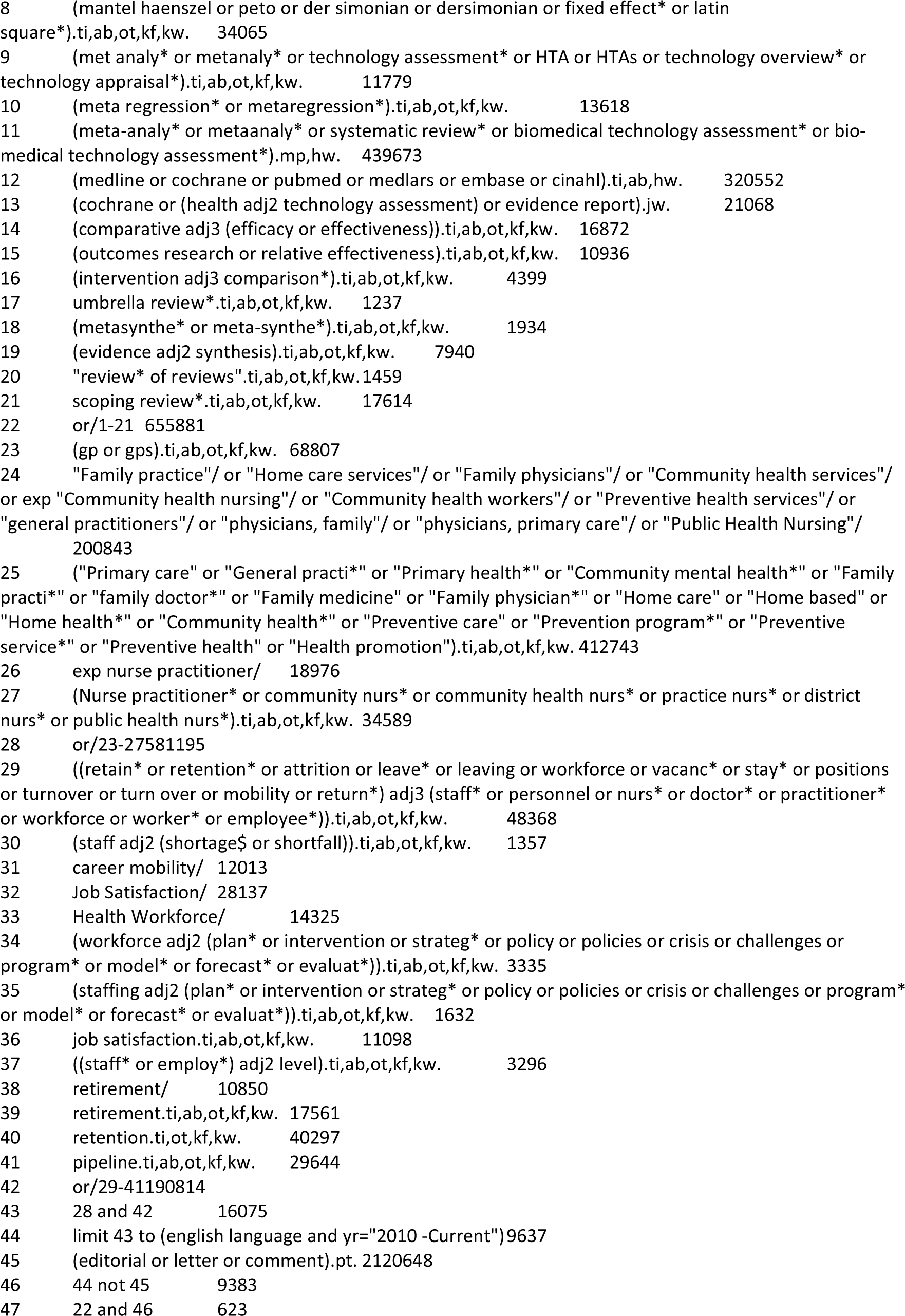
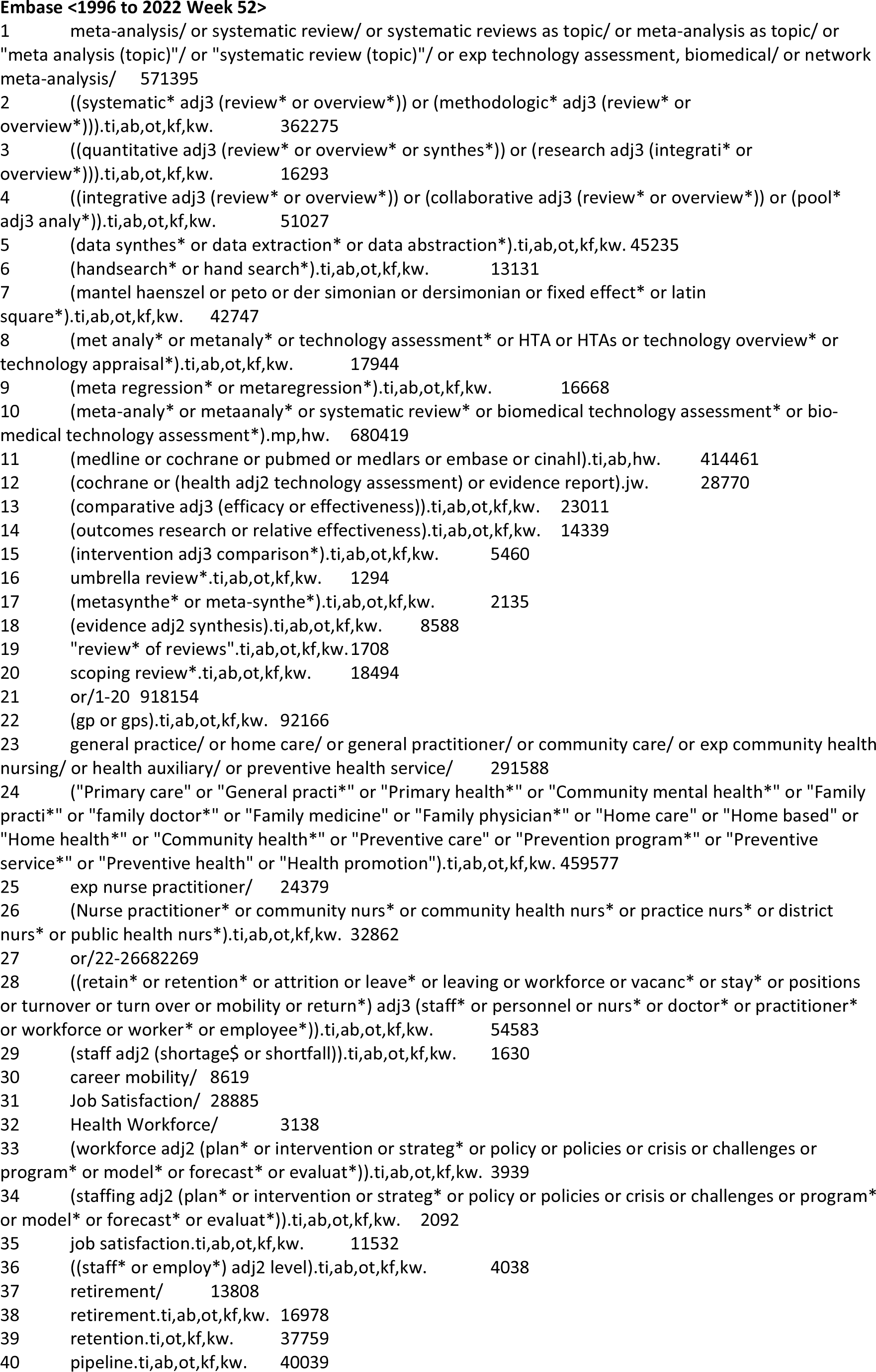
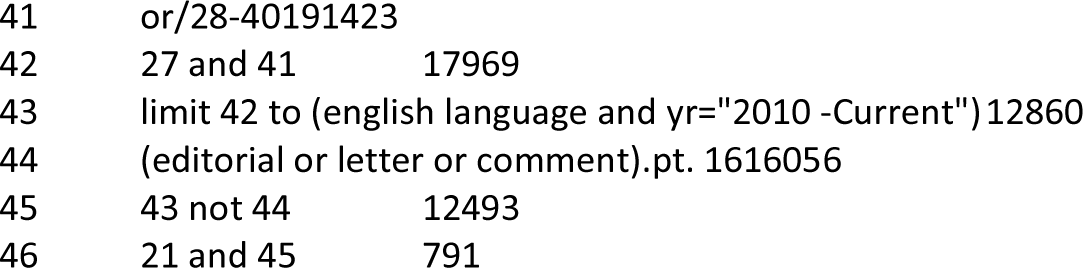
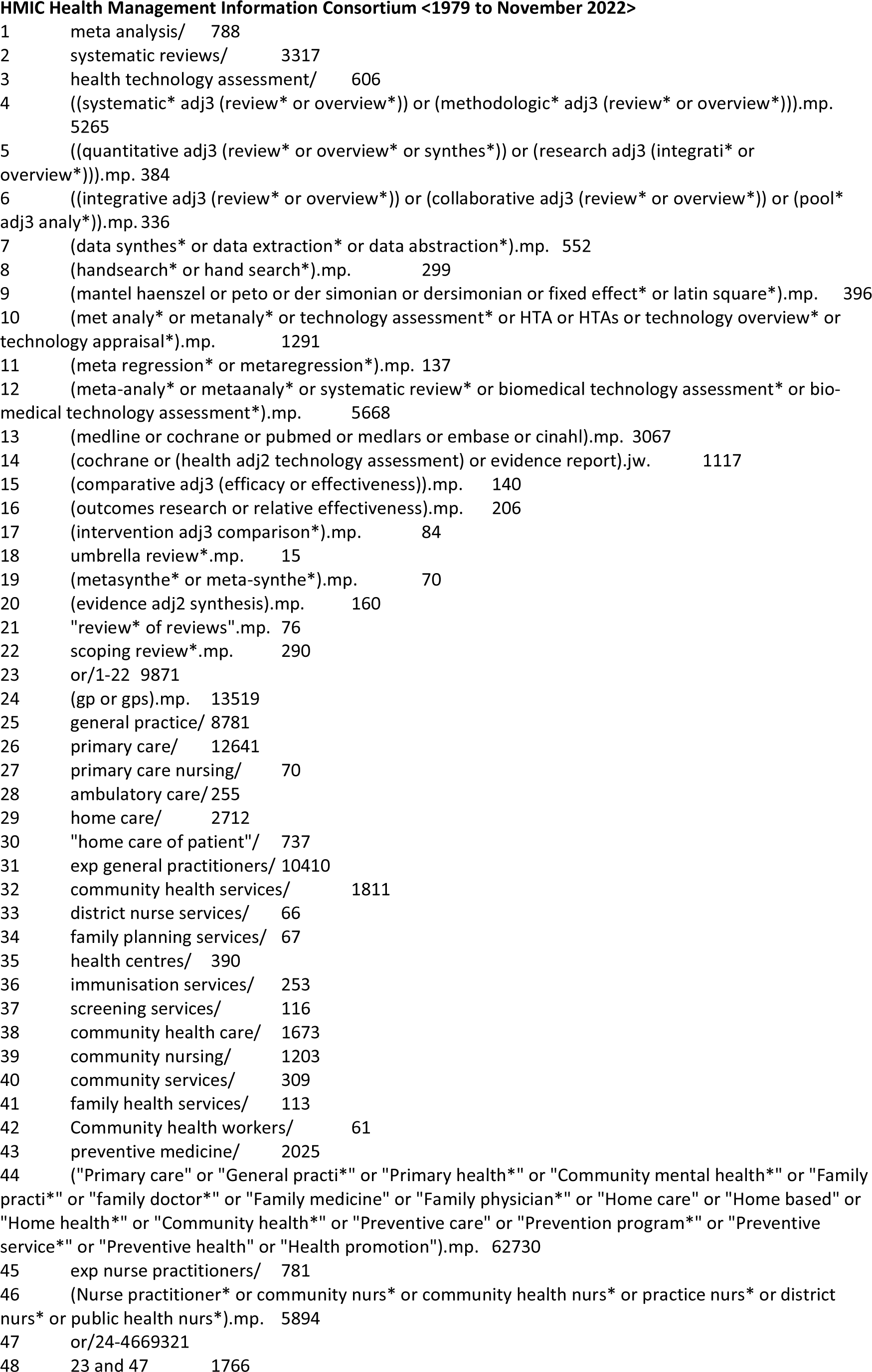
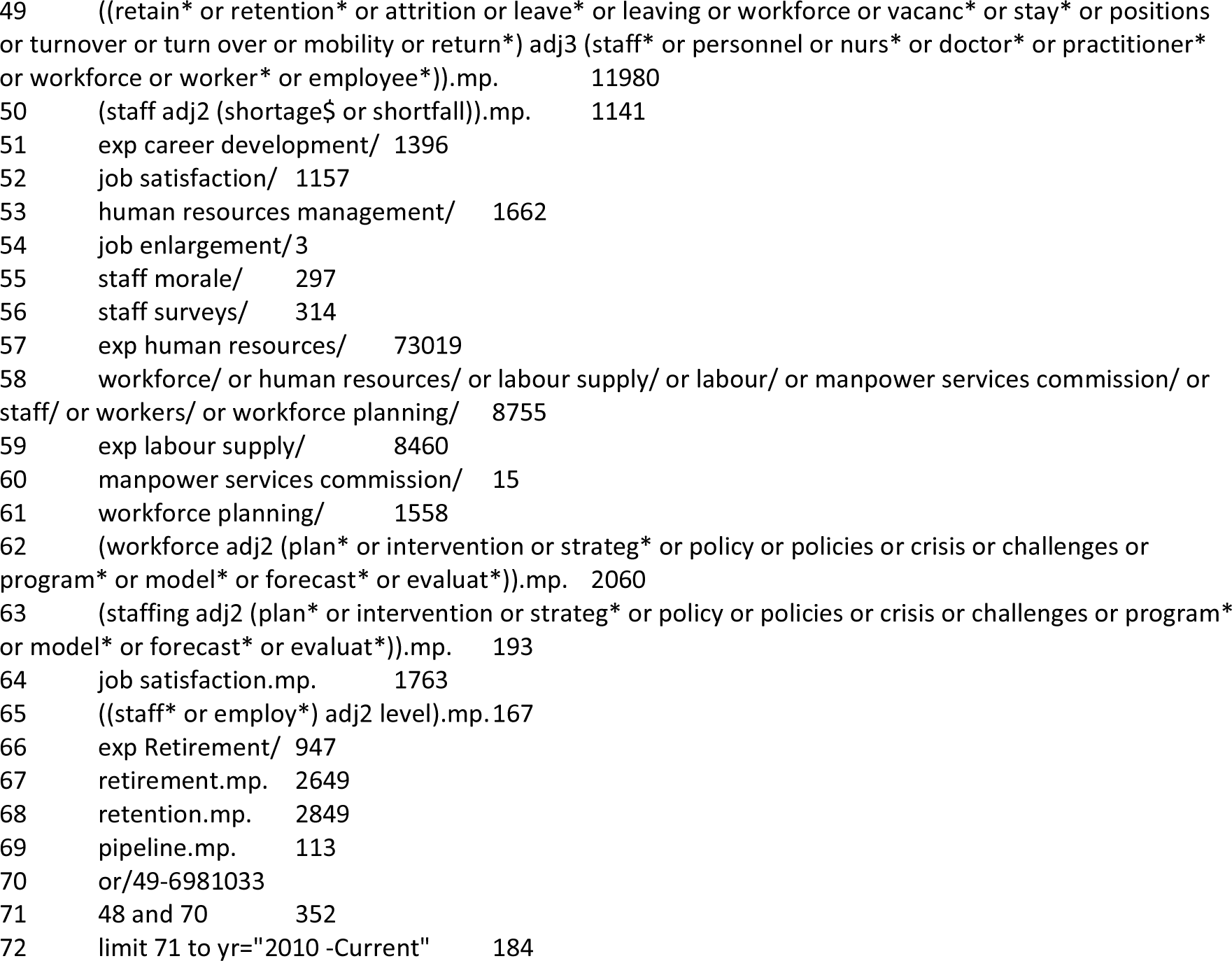

### CINAHL Complete

((PT “systematic review”) OR (PT meta-analysis)) OR (MH “systematic review”) OR ((TI review*) OR (AB “systematic review*”) OR (TI “systematic overview*” OR AB “systematic overview*”) OR (TI “evidence synthes*” OR AB “evidence synthes*”) OR (TI “data synthes*” OR AB “data synthes*”) OR (TI “data extraction*” OR AB “data extraction*”) OR (TI “data abstraction*” OR AB “data abstraction*”) OR (TI handsearch* OR AB handsearch*) OR (TI “hand search*” OR AB “hand search*”) OR ((TI “mantel haenszel” OR AB “mantel haenszel”) OR (TI peto OR AB peto) OR (TI “der simonian” OR AB “der simonian”) OR (TI dersimonian OR AB dersimonian) OR (TI “fixed effect*” OR AB “fixed effect*”) OR (TI “latin square*” OR AB “latin square*”) OR (TI “meta analy*” OR AB “meta analy*”) OR (TI metaanaly* OR AB metaanaly*) OR (TI “technology assessment*” OR AB “technology assessment*”) OR (TI HTA OR AB HTA) OR (TI HTAs OR AB HTAs) OR (TI “technology overview*” OR AB “technology overview*”) OR (TI “technology appraisal*” OR AB “technology appraisal*”) OR (TI “meta regression*” OR AB “meta regression*”) OR (TI metaregression* OR AB metaregression*) OR (TI medline OR AB medline) OR (TI cochrane OR AB cochrane) OR (TI pubmed OR AB pubmed) OR (TI medlars OR AB medlars) OR (TI embase OR AB embase) OR (TI cinahl OR AB cinahl) OR (AB “umbrella review*”) OR (TI metasynthe* OR AB metasynthe*) OR (TI meta-synthe* OR AB meta-synthe*) OR (AB “review of reviews”) OR (AB “scoping review*”))

### AND

((TI gp OR AB gp) OR (TI gps OR AB gps) OR (MH “Primary Health Care”) OR (MH “Family practice”) OR (MH “Community health services”) OR (MH “Family Nurse Practitioners”) OR (MH “Community health nursing+”) OR (MH “Community health workers”) OR (MH “physicians, family”) OR (TI “Primary care” OR AB “Primary care”) OR (TI “General practi*” OR AB “General practi*”) OR (TI “Primary health*” OR AB “Primary health*”) OR (TI “Community mental health*” OR AB “Community mental health*”) OR (TI “Family practi*” OR AB “Family practi*”) OR (TI “family doctor*” OR AB “family doctor*”) OR (TI “Family medicine” OR AB “Family medicine”) OR (TI “Family physician*” OR AB “Family physician*”) OR (TI “Nurse practitioner*” OR AB “Nurse practitioner*”) OR (TI “community nurs*” OR AB “community nurs*”) OR (TI “community health nurs*” OR AB “community health nurs*”) OR (TI “practice nurs*” OR AB “practice nurs*”) OR (TI “district nurs*” OR AB “district nurs*”) OR (TI “public health nurs*” OR AB “public health nurs*”))

### AND

(((TI retain* OR AB retain*) OR (TI retention* OR AB retention*) OR (TI attrition OR AB attrition) OR (TI leave* OR AB leave*) OR (TI leaving OR AB leaving) OR (TI workforce OR AB workforce) OR (TI vacanc* OR AB vacanc*) OR (TI stay*) OR (TI positions) OR (TI turnover OR AB turnover) OR (TI “turn over” OR AB “turn over”) OR (TI career OR AB “career mobility” OR AB “career progession”) OR (TI staff* OR AB “staff* level*”) OR (TI personnel) OR (TI employee* OR AB employee*) OR (TI pipeline OR AB pipeline) OR (TI “job satisfaction” OR AB “job satisfaction”) OR (TI “retirement” OR AB “retirement”))) OR (MH “Personnel Retention”) OR (MH “Personnel Shortage”) OR (MH retirement) OR (MH “career mobility”)

Limit to English language and publication date 2010 onwards

### British Nursing Index (BNI)

“systematic review*” OR “systematic overview*” OR “evidence synthes*” OR “data synthes*” OR “data extraction*” OR “data abstraction*” OR handsearch* OR “hand search*” OR “mantel haenszel” OR peto O “der simonian” ORI dersimonian OR “fixed effect*” OR “latin square*” OR “meta analy*” OR metaanaly* OR “technology assessment*” OR HTA OR HTAs OR “technology overview*” OR “technology appraisal*” OR “meta regression*” OR metaregression* OR medline OR cochrane OR pubmed OR medlars OR embase OR cinahl OR “umbrella review*” OR metasynthe* OR meta-synthe* OR “review of reviews” OR “scoping review*”

### AND

gp OR gps OR “Family practice” OR “Community health services” OR “Community health nursing” OR “Community health workers” OR “physicians, family” OR “Primary care” OR “General practi*” OR “Primary health*” OR “Community mental health*” OR “Family practi*” OR “family doctor” OR “Family medicine” OR “Family physician*” OR “Nurse practitioner*” OR “community nurs*” OR “community health nurs*” OR “practice nurs*” OR “district nurs*” OR “public health nurs*”

### AND

Retain OR retention* OR attrition OR leave* OR leaving OR workforce OR vacanc* OR turnover OR “turn over” OR career OR “staff* level*” OR “staff shortage*” OR “staff plan*” OR employee* OR pipeline OR “job satisfaction” OR retirement

Limit to publication date 2010 onwards

## Data Availability

Data is available on request from study authors.

